# Drivers and barriers to the implementation of the school feeding values-based food procurement guidelines and ultra-processed food restrictions

**DOI:** 10.64898/2026.04.22.26351508

**Authors:** Vanessa Fernandes Davies, Igor Perrut, Anne-Marie Thow, Ana Clara Duran

**Author notes:** Corresponding author: Ana Clara Duran.

## Abstract

**Objective:** To investigate in the National School Feeding Program (PNAE) the local-level drivers and barriers to the implementation of four guidelines: the banning of sugary drinks; restrictions on the procurement of processed and ultra-processed foods; the mandatory increase in weekly servings of fruits and vegetables offered to students; and mandatory direct procurement from family farmers.

**Design:** Qualitative study that used semi-structured interviews. Street-level bureaucracy theory informed the theoretical framework and thematic analysis.

**Setting:** Brazilian municipalities, across the country’s five geographic regions (North, Northeast, Southeast, South, and Midwest).

**Participants:** Stakeholders (e.g. nutritionists, school cooks, and food procurement managers) involved in the local implementation of the PNAE program across the country.

**Results:** Ninety stakeholders were interviewed. Stakeholders reported having autonomy to perform their activities, collaboration and support from other members within the local government and food providers, adequate infrastructure such as a well-equipped kitchens, the availability of trained personnel, and political commitment as drivers for optimum program implementation. Reported barriers included lack of support and resistance to change among cooks, teachers and parents; insufficient physical and human resources; and limited political commitment. When barriers outweighed drivers, interviewees reported adapting their practices, often in restrictive ways that could compromise the implementation of the program.

**Conclusions:** Drivers and barriers to local PNAE implementation were generally similar across studied municipalities, although their magnitude varied. In contexts of greater economic vulnerability and fiscal constraint, additional support and targeted actions from the federal government may be required to strengthen local implementation.

## Introduction

The National School Feeding Program (PNAE) in Brazil is one of the largest school feeding programs in the world. Approximately 50 million meals are produced daily, serving 40 million students in 150,000 Brazilian schools^1^. The program has served as a reference for the development of school feeding initiatives in other countries, contributing to the implementation of programs in 29 countries in Latin America and the Caribbean through the Brazil–FAO Trilateral South–South Cooperation Project, and supporting projects of the World Food Programme’s Centre of Excellence against Hunger in 28 countries across the African continent^1^.

PNAE was created in the 1950s with the aim of helping to reduce hunger at a time when the Brazilian population faced very poor health and nutritional conditions^2^. Since then, alongside the demographic, nutritional, and epidemiological changes experienced by the Brazilian population^3,4^, the program has undergone several important developments. These include the expansion of program coverage, increased allocation of financial resources, the strengthening of technical and operational criteria, the expansion and institutionalization of social control mechanisms, and the establishment of the Collaborating Centers for School Food and Nutrition (CECANE)^5^.

In 2009, Law No. 11.947^6^ was enacted, stipulating that at least 30% of the program’s funds must be allocated to the direct procurement of products from family farmers. This provision has contributed to stimulating the sustainable economic development of local communities^1–7–10^.

Currently, PNAE plays a fundamental role in promoting food and nutrition security among the Brazilian population^11^, while also contributing to the livelihoods of smallholder farmers^12^. From a nutritional perspective, the program both promotes the provision of healthy foods and restricts the availability of unhealthy products offered to students. These measures include the legal prohibition of sugary beverages in schools, and a limit of 20% of federal program funds that may be used for the procurement of processed or ultra-processed foods^13,14^. In addition, Resolutions No. 26 of June 2013 and No. 6 of May 2020 establish minimum weekly requirements for the provision of fruits and vegetables in school meals^14–16^.

The planning and implementation of PNAE involve the participation of a network of diverse stakeholders and collaborators^11^. This network includes nutritionists (who are legally responsible for developing school meal menus), professionals involved in public food procurement, cooks, and other support staff. It also involves institutional partners such as the School Food and Nutrition Collaborating Centers (CECANE), which provide technical support and training to PNAE stakeholders, as well as civil society representatives such as the School Food Council (CAE), whose role is primarily to monitor and oversee program implementation. In addition, family farmers and the organizations that support them are key actors in the stakeholder network, particularly in relation to the implementation of the family farming procurement mandate established under Law No. 11.947^6^.

Existing studies evaluating PNAE implementation have largely focused on specific actors or municipalities within a single region of Brazil, limiting understanding of how the program operates across diverse local contexts. As a result, there remains limited evidence of the factors that facilitate or hinder the implementation of its key nutrition and procurement guidelines at the municipal level. To address this gap, this study draws on a national sample of diverse PNAE stakeholders from 15 municipalities across Brazil. The objective of this paper is to examine the local-level drivers and barriers to the implementation of PNAE’s values-based food procurement guidelines and restrictions on ultra-processed foods.

## Methods

### Study Design

This qualitative study used semi-structured interviews to analyse the implementation of Law No. 11.947 (2009), and the Resolutions No. 26 of June 2013 and No. 6 of May 2020 of Brazil’s National School Feeding Program (PNAE).

Lipsky’s theory of ‘Street-Level Bureaucracy’ (2010)^17^ was used as a theoretical framework to guide the fieldwork, plan the data collection tools, and analyze the results.

The authors chose Lipsky’s theory^17^ to understand how the actors involved in implementing the PNAE deal with the different demands and pressures that they experience when implementing the laws evaluated here. Lipsky’s theory^17^ considers that those responsible for implementing laws, policies, and programs for citizens use discretionary practices, which are ways of carrying out their work within the inherent limits of public service (e.g., limitations of financial and/or human resources). In other words, they adapt their work to the context, translating the theory to the day-to-day reality of public services.

### Study Setting and Participants

The study used the principle of outlier sampling^18^. The objective was to include municipalities that demonstrated both high and low levels of compliance detailed and descriptive information on how the implementation of the National School Feeding Program (PNAE) occurs.

We used publicly available information from the National Accountability System reports from the National Fund for the Development of Education (FNDE)^19^, and from the Brazilian Ministry of Education^1^ to select the municipalities for study. All Brazilian municipalities are required to report the food items procured using federal transfers allocated to the National School Feeding Program (PNAE).

We used data from 2019, the most recent year available prior to the disruptions caused by the COVID-19 pandemic, during which schools were closed for extended periods.

All procured food items were cleaned, treated, and classified according to:

1. The procurement modality—whether purchases were made through standard public bidding processes or through direct procurement from family farmers; and
2. The degree of food processing, based on the NOVA food classification system.

Using these classifications, municipalities were categorized based on (i) the percentage of federal transfers used to procure food directly from family farmers, and (ii) the percentage of federal transfers used to procure processed and ultra-processed foods.

Based on these indicators, municipalities were grouped (Figure 1).

**Figure 1:**
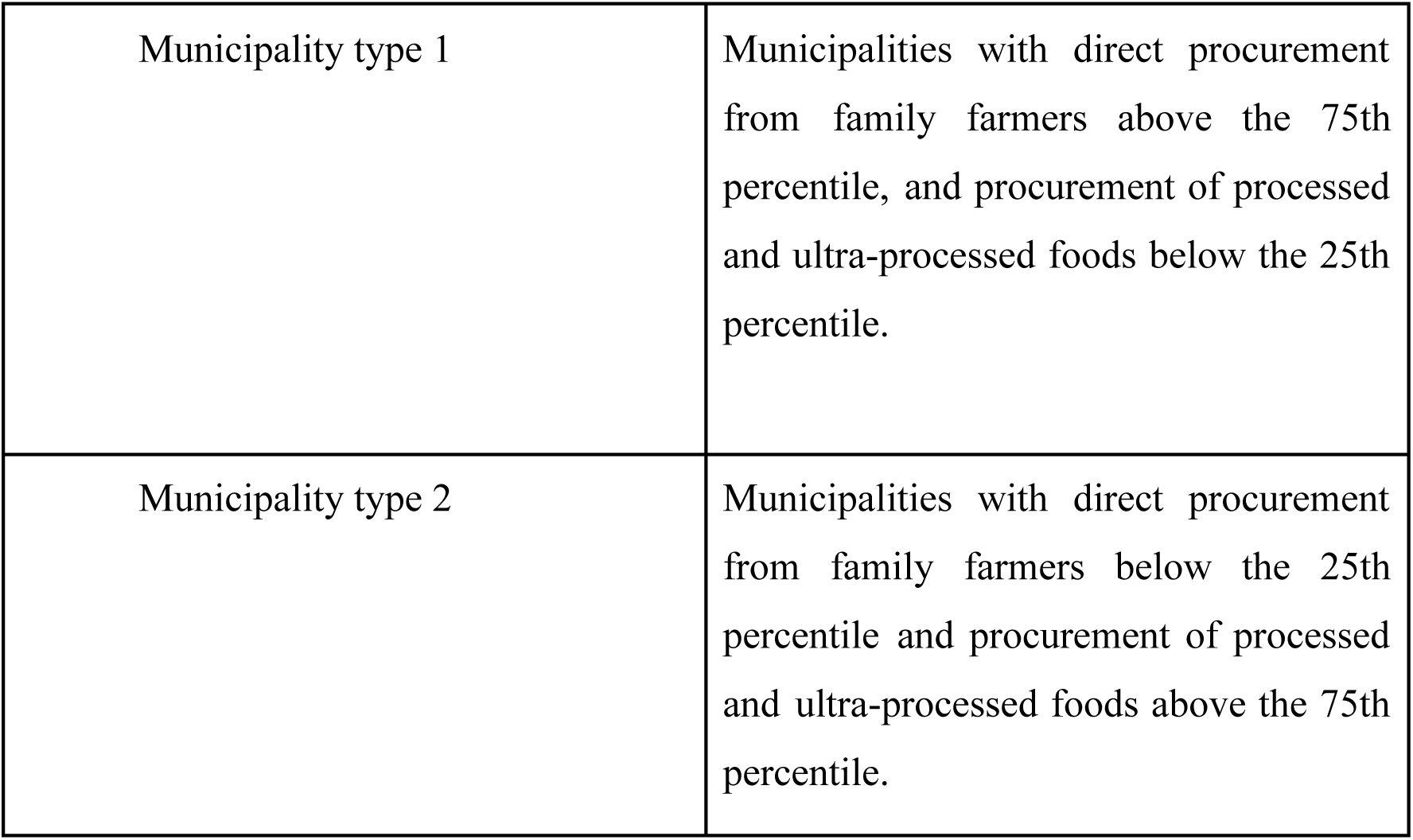
Classification of municipalities according to Federal transfers to purchase food.

Municipalities were purposively sampled within these groups to ensure representation from all the geographic regions of Brazil. The final sample included fifteen municipalities distributed across the country’s five macro-regions, encompassing both urban and rural settings.

Municipalities were initially invited to participate through telephone contact. Subsequently, letters introducing the study, along with acceptance forms, were sent to the Municipal Secretariats of Education for formal approval. Each municipality was then asked to identify the nutritionist responsible for the local implementation of the school feeding program.

The nutritionist was selected as the first interviewee in each municipality, because their menu-planning activities initiate and coordinate several key processes related to the implementation of the regulations evaluated in this study. In addition, nutritionists assisted the research team in identifying and providing contact information for other relevant participants through snowball sampling techniques.

Interviewees were selected based on their roles within PNAE and were organized into three groups (Figure 2).

**Figure 2:**
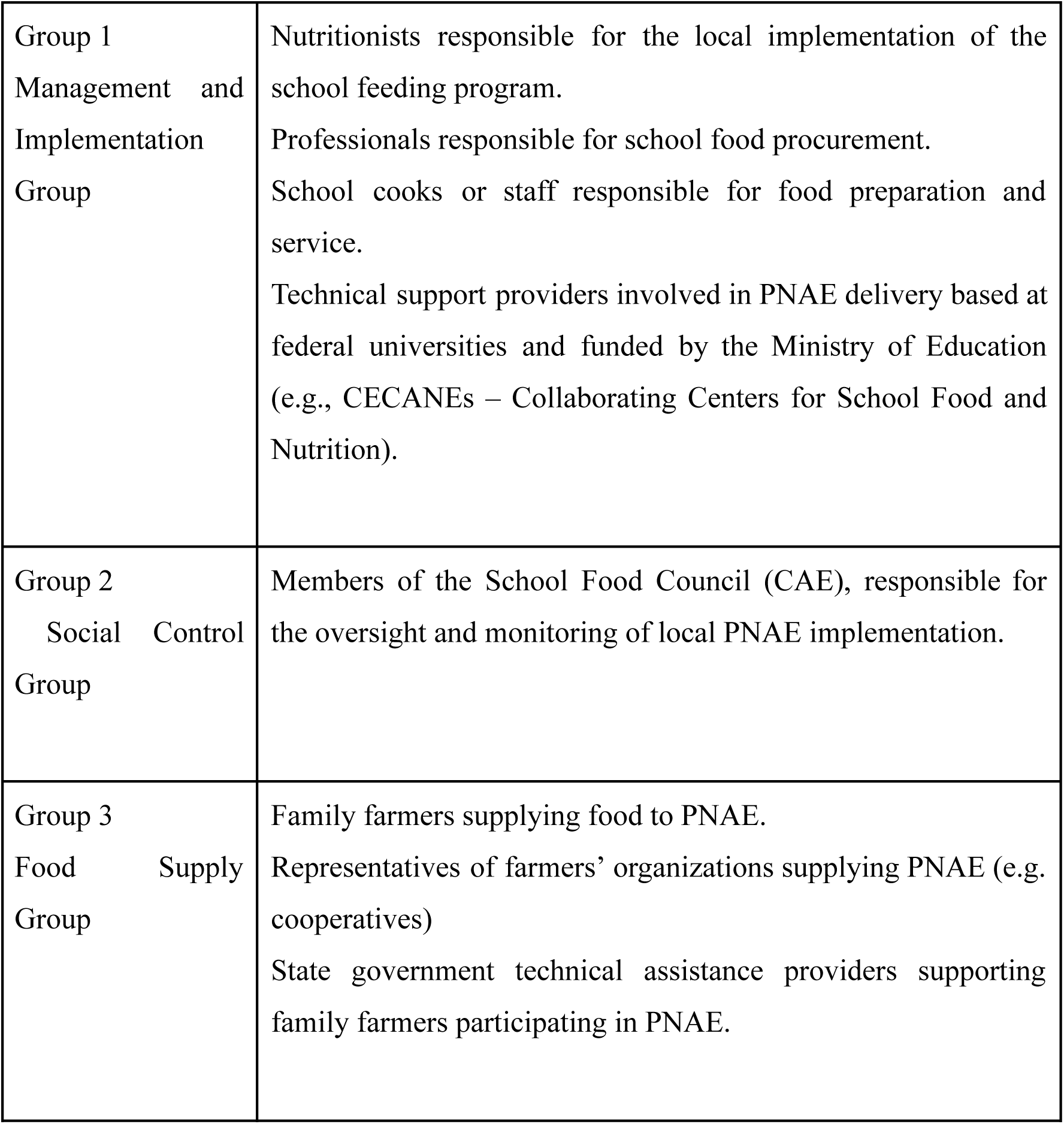
Distribution of PNAE actors by group.

### Data Collection

A semi-structured interview technique was used, adopting a storytelling approach to ensure that the interview flow remained natural and comfortable for participants while maintaining a conversational atmosphere and focusing on the research objectives. Interviews were conducted in Portuguese by researchers trained in qualitative research methods.

Individual interview guides were developed for each participant group, and were based on the study’s main research question. The guides included: (i) an opening question to understand the interviewee’s daily activities and role in the program; (ii) core questions aimed at understanding how the regulations evaluated in this study are implemented in practice; and (iii) a closing question allowing participants to raise additional issues they considered relevant but that had not been addressed during the interview. For each main question, follow-up probes were prepared to further explore and expand participants’ responses.

The interview guides were reviewed and tested by members of the research team. In addition, the guide designed for nutritionists was pilot-tested in two municipalities that were not included in the study sample. This process aimed both to assess the suitability of the questions for addressing the research objectives, and to familiarize the researchers responsible for conducting the interviews with the interview protocol.

Most interviews were conducted online using the Google Meet platform, and were audio-recorded with participants’ consent for subsequent transcription. Interviews lasted approximately one hour and thirty minutes. In cases where participants were unable to access Google Meet, interviews were conducted via WhatsApp as an alternative form of communication.

### Data Analysis

Interview recordings were transcribed using NVivo 14 software (2025)^20^, and subsequently reviewed by members of the research team to ensure transcription accuracy.

Data were analyzed using thematic analysis following the approach described by Braun and Clarke^21^ (2006), combining both deductive and inductive analytical strategies. The deductive component involved allocating data to predefined themes derived from the theoretical framework and relevant literature, while the inductive component allowed new themes to emerge from the empirical material. Below we describe the main steps in the analytical process.

For the deductive phase, we conducted literature reviews and examined preliminary interview data to better understand how discretion operates in the implementation of public policies. Based on these insights, the research team held a series of analytical meetings to define the initial thematic categories for reporting how PNAE actors interpret and operationalize program regulations in their daily practices. These themes particularly focused on the routines during which actors simplify and adapt policy requirements, as well as the dynamics shaping local implementation processes. Data analysis was conducted in Portuguese, and the results were subsequently translated into English to allow all co-authors to review and contribute to the interpretation of the findings. The themes developed for the deductive stage of the analysis are shown in Table 1.

**Table 1.** Predefined codes for analyzing the routines and dynamics of implementing the laws evaluated (Codebook).

| Topics | Description |
| --- | --- |
| Discretionary decisions | Issues related to choices and decisions made by the implementers in order to perform their work, such as the creation of alliances or support coalitions, construction of political-institutional matrices of cooperation or conflict, decision-making based on their own understanding about how their work should be done, or actions related to the challenging points that are beyond their control, knowledge of the program, agreement (or lack thereof) with the program's objectives. |
| Institutional activism | Content related to the actors' moral views of the implementing the PNAE, such as values, opinions, motivations, interests, perspectives, beliefs, intentions, desires, and ideological views. |
| Accountability | Topics that are related to the relationship between implementers and clients, inquiring into the networks of accountability, criticism, referencing and ways of support for the public policy assistance. |
| Context | Issues related to the financial resources available for the program implementation, pressures for decision-making, internal and administrative responsibilities, availability of human resources to support implementation. |
| Policy design | Social, political, economic and cultural issues that influence the local and contextual implementation of the program. |

Regarding the inductive phase of the analysis, NVivo 14 software (2025)^20^ was used to organize the collected data and support the analytical process. The coding and analysis were conducted independently by two researchers.

Following the individual analyses, the researchers met to compare and consolidate their findings. Any discrepancies in coding or interpretation were discussed until consensus was reached.

The analytical process consisted of the following steps: (i) familiarization with the data, which included reading and rereading the transcripts;(ii) coding extracts for the topics presented in the codebook (see Table 1); (iii) from reading the extracts allocated to the codebook (pre-established topics), the process of generating codes was redone; (iv) searches for inductive codes were carried out; (v) themes were created, revised and named in order to create thematic maps for each actor and each group of actors. (vi) the similarities and discrepancies within each theme were discussed among the researchers, and the content of each theme was refined according to how it related to the research question. Finally, (vii) thematic maps were verified against the principle of homogeneity (i.e. did they all focus on the same theme?) and the principle of heterogeneity (i.e. is each theme different from the others?).

The final thematic map illustrates the drivers and barriers reported by interviewees across key stages of program implementation, including menu planning, food procurement, interactions with suppliers and technical support staff, and the preparation and distribution of school meals. The thematic maps generated through the analysis are provided in the supplementary material.

The final thematic map demonstrates the drivers and barriers encountered by the interviewees throughout the stages of menu planning, food purchasing, relationships with suppliers and technical support staff, as well as the preparation and distribution of school meals. The thematic maps created are available in the supplementary material.

## Results

Initially, 120 people were selected to participate, seven people did not answer any form of interview invitation, five people declined to participáte and three people had no contact information to send invitations. Overall, the study completed 75% of the planned interviews. Details of the sample can be seen in Table 2.

**Table 2.** Distribution of participants by group and region.

| **Actors/**<br>**Region** | **North\*** | **North**<br>**East** | **Midwest**<br>**\*** | **Southeas**<br>**t** | **South** | **Total**<br>**Group** |
| --- | --- | --- | --- | --- | --- | --- |
| Group 1 | 8 | 14 | 7 | 11 | 10 | 50 |
| Group 2 | 2 | 4 | 2 | 4 | 3 | 15 |
| Group 3 | 3 | 8 | 3 | 5 | 6 | 25 |

\* The North and Midwest regions, which are the least populated regions of Brazil, also contributed fewer municipalities to the study sample.

The results of the thematic analysis were organized into two main themes—drivers and barriers—and their corresponding subthemes (Table 3).

**Table 3.** Final main themes and subthemes with their descriptions originated from the analyses.

| Theme | Subthemes | Description of subthemes |
| --- | --- | --- |
| Drivers | Ways of doing | Names the set of actions that are seen as external to those prescribed in the resolutions, but without which they themselves could not be implemented. |
|  | Actors' Network | Refers to the strengthening of close relationships built between program actors, using them as an element that facilitates program implementation. In addition, the availability of partnerships that help to improve knowledge among people who participate in PNAE directly or indirectly. |
|  | Human Resources and Infrastructure | All elements that provide the foundation upon which the program operates, including physical operational structures—such as inventory, kitchen appliances and |
|  |  | workspaces, financial resources, and the working conditions of PNAE actors. |
|  | Political commitment | Actions that delimit the implementation of a political environment favorable to increased food spending, as well as the use and investment in school meals and the implementation of the laws evaluated in this study. |
| Barriers | Financial constraints | Includes the factors that are related to a lack of financial resources to ensure the implementation of the PNAE. |
|  | Lack of partnership or communication | Actors' dynamics that are absent to promote dialogue, support and conflict resolution. |
|  | Food system challenges | Issues related to family farmers' workload, agricultural challenges and climatic adversity. |
|  | Lack of political commitment | Political environments that are not aligned with the PNAE objectives, or have |
|  |  | vested interests in the program. |
|  | Resistance to Change | Obstacles created to prevent or, at the very least, resist changes to the menu and its implementation. |

Drivers and barriers will be detailed below, and examples of perspectives from actors for each theme can be seen in Table 4.

**Table 4.** Drivers and barriers according to groups and related examples of perspectives from participants.

| Groups that generated the themes | Drivers | Example of perspectives related to drivers | Barriers | Example of perspectives related to barriers |
| --- | --- | --- | --- | --- |
| Group 1 (Nutritionists; actor responsible for purchasing school meals; Cooks; and Collaborating Centers for School Food and Nutrition - CECANE). | Ways of doing: Autonomy and being proactive. | *Yes, because here I have a lot of autonomy. So, my superiors—the mayor, deputy mayor, education secretary, and everyone else—they let me actually organize the meals... the school meals are in my hands. (Nutritionist)* | Food system: Issues related to family farmers' limited resources or inability to supply the demand from PNAE. | *So, I think it's even a matter of their own production, right? Because I wanted to ask for more pineapple. And then the producers said they didn't have enough. I think they need to increase their production, right? (Nutritionist)* |
| Group 1 (Nutritionists; responsible for purchasing school meals; Cooks; and Collaborating Centers for School Food and Nutrition - CECANE) + | Actors network: communication and training. | *So, when we work together with the school and start providing information, I believe it's more successful. Difficulties are reduced* | Lack of partnership or communication: limited knowledge about what food is available in the region, and confusion | *The first thing you need to know is what you have in your region and what is produced and what isn't, so the nutritionist can decide whether or not to* |
| Group 3 (Family farmers; representatives of farmers' organizations (cooperatives); and actors responsible for providing technical support to family farmers supplying the PNAE). |  | *because when we work together, we inform each other and provide assistance, and everyone supports each other. (Social control)*<br>*The manager, nutritionist, School Nutrition Council, Health Department... Everyone has built a climate of trust among themselves. This intersectoral trust is what I'm talking about. (CECANE)* | about the process of selling food to PNAE | *include it on the menu. And we always ask: is there an agricultural map of the region? There never is.*<br>*(Actor responsible for providing technical support to family farmers supplying the PNAE)* |
| Group 1 (Nutritionists; responsible for purchasing school meals; Cooks; and Collaborating Centers for | Human resources and Infrastructure:<br>Enough municipal financial investment to | *Look at this mayor! He really encourages this group of farmers. He doesn't sit at home and do* | Municipalities with financial constraints: not hiring enough people to implement the PNAE and | *Given the number of cooks we have at my school, sometimes it leaves something to be* |
| School Food and Nutrition - CECANE) + Group 2 (Social Control). | plan, prepare and deliver healthy meals and economic support to develop the family farming in the region. | *nothing. He goes to the rural area, holds meetings with the producers, and gives them seeds, you know? In our region, we didn't have carrots, so he hired agronomists and gave them carrot seeds...*<br>(Actor responsible for purchasing school meals)<br>*This current administration in charge of local government has welcomed children's nutrition and has equipped all the schools. They have provided them with equipment. So, while*<br>*you're asking to buy a refrigerator; the manager never refused, or said they don't have the money for it. (Social Control)* | lack of infrastructure to produce and store food at schools. | *desired, because we don't always have time to take the necessary care, to make everything nice, and sometimes we don't have time... We're at the school where I work now, with 450 children. There's just me and one other cook.*<br>(Cook) |
| Group 1 (Nutritionists; Actors responsible for purchasing school meals; Cooks; and Collaborating Centers for School Food and Nutrition - CECANE) + Group 2 (School Food Council - CAE). | Political commitment to comply with the laws | *In municipalities where the very basis of the menu is natural, with minimally processed foods, that provide food and nutrition education, that buy from family farms, there are usually people behind them who support them: the mayor, the nutritionist who is closely involved and has a voice. (CECANE)* | Lack of political commitment | *In both urban and rural schools, we still see administrators buying prohibited foods that are listed in resolution number six, and sometimes the nutritionist sends the purchasing food list. So what? And then the department that conducts the public procurement process buys differently, doesn't buy what's recommended for a healthy diet, and they*<br>*know they have to comply with the law. And they'll also be monitored by the PNAE and held accountable. But they also face issues of business interests, where some administrators may have ties to suppliers, and therefore their hands are often tied.*<br>(CECANE) |
| Group 1 (Nutritionists; responsible for purchasing school meals; Cooks; and Collaborating Centers for School Food and Nutrition - CECANE) + Group 2 (Social Control group) | - | - | Resistance to change | *The school staff ends up having a lot of influence on both acceptance and rejection. So, this year we had a meeting at a school where, talking to the teachers, the following statement was made:* |
|  |  |  |  | *"Because I don't do it that way in my house—but it's not your house!" Most of the time, the nutritionist needs to be assertive and demonstrate their work. So, the food is only for the student. The teacher is there to encourage, to monitor, but if they don't realize the importance, it ends up slightly hindering the acceptance of food.*<br>(Nutritionist) |

### Drivers

A sense of autonomy was perceived by the nutritionists as something that helped to implement the PNAE. Examples of what the word autonomy meant in PNAE daily activities included: creating a menu without managerial interference; the absence of financial constraints when choosing and purchasing food; and having the knowledge and experience to design and prepare healthier menus. The autonomy to comply with the laws assessed here, according to the interviewees, was achieved mainly through the hard work of the nutritionist trying to help managers understand the importance of the laws. In addition, it was also reported that nutritionists who had the security of being civil servants (i.e. they cannot be dismissed from their positions) favored a more autonomous approach to implementing the laws.

Based on the awareness of the importance of providing healthy food for children, additional activities (beyond their usual duties) were performed by some actors, such as: school cooks being proactive in creating recipes and working to convince students of the importance of healthy eating; family farmers delivering food door-to-door to schools that lack a central collection point; principals prohibiting students from bringing unhealthy foods to school; and Social Control bodies conducting food and nutrition education activities in schools.

It was also understood by the participants that building relationships with all the actors is an important driving factor for the PNAE implementation. In the process of planning the menu for the schools, the nutritionist needed to gain knowledge about food produced in the region, and together with other actors (for example producers and cooks), to work on problem-solving situations such as delivery delays, food replacements, excess bureaucracy, unforeseen weather conditions, and other factors that impact the day-to-day running of the PNAE. In this regard, the fact that the nutritionist could count on partnerships with external organizations, helped to reassure them that the planned menu would be transformed into real food at schools.

Technical support actors (Group 3 - linked to family farmers, and family farmer cooperatives) were particularly noteworthy as an external partnership, providing training for family farmers on topics related to “what and when to plant,” possibilities for innovation in the production chain, among other things. The Cecane (Group 1 - management & implementation) was another support group crucial for the nutritionists, helping to mediate relationships and training stakeholders in the PNAE legal compliance. It was mentioned by the participants that in order for the network of actors of PNAE to work properly, everyone needs to understand how the laws of the program work, and to be trained accordingly, from those who purchase food, suppliers and inspectors, to those who deal with children in schools such as cooks, teachers and principals. The role of Cecane was highlighted as a fundamental external actor, not only helping to identify the gaps in the program for administrators, but also throughout the entire PNAE activity network, developing support materials and conduct training workshops on “how, what, and why to do it” with all stakeholders, raising awareness and providing training on adequate and healthy nutrition in schools.

Within the scope of human resources and infrastructure, the analyses indicated that having an adequate number of professionals in each role helped to guarantee that the PNAE actors could perform their activities in an appropriate way, such as making sure that the foods served at school followed the planned menu designed by the nutritionists. Well-equipped kitchens (i.e. investment in the infrastructure) with apparatus for preparing and storing food was also reported as being important to be able to produce more elaborate recipes, optimize their kitchen activities, and avoid food waste.

Aspects related to administrators’ awareness and commitment to complying with PNAE laws demonstrated that when there is a commitment within the city government to comply with the laws, the network of activities within the PNAE operates more smoothly. For example, the value placed on family farming by the municipality encouraged activities to increase the availability of family farmers in the region, as well as more municipal investment in developing and attracting family farmers in regions where these actors are scarce. The allocation of substantial amounts of municipal resources for purchases from family farmers in some municipalities was perceived as an incentive for suppliers to grow a larger and more varied quantity of food to meet the PNAE’s needs. In addition, there was a greater concern with oversight, fostering openness for Social Control members to carry out their work, and accepting recommendations for PNAE improvements from CECANE.

### Barriers

In municipalities with financial constraints on purchasing PNAE food, stakeholders have greater difficulty developing and implementing healthier menus that follow PNAÉs guidelines to restrict processed and ultra-processed foods (Brasil, 2020, Canella et al., 2021). It is noteworthy that in these situations, it was reported that cheaper foods may be included on menus, regardless of whether students like them, and the quantity of fruit offered may be low, failing to meet recommended levels. Issues such as late payments and dependence on federal funding were mentioned, due to the financial instability of some regions. Difficult situations in which there are too few professionals to properly implement the PNAE, for example in schools that serve a large number of meals per day, creating an overload of work for cooks; and municipalities with numerous schools spread across urban and rural areas which can experience obstacles to monitoring the implementation of the laws by the nutritionists or CAE members.

Furthermore, there still appears to be a mismatch between planned menus and what is produced locally. On the one hand, the actors responsible for planning and buying the food for the school meals appeared not to have an agricultural map of local suppliers (i.e. not knowing who the family farmers available in the area were) or to have any kind of partnership with family farmers. In addition, problems related to the infrastructure of schools, such as a lack of food storage facilities (either at a central location or at each school), resulted in financial losses for farmers, impacted their participation in the PNAE, and also affected the variety and quantity of fruit distributed to students. On the other hand, food system barriers such as: a lack of farmers in the region; a limited variety of fruits and vegetables produced; problems for farmers to produce adequate quantities to meet the PNAE’s needs; delivery delays; and climate issues which negatively impacted the PNAE actors’ work.

It was frequently reported that the successful implementation of laws appears to be dependent on those in positions of power in the municipalities. In other words, those in power may view school meals as a form of investment in education, or simply as an obligation to be fulfilled. As a result, the role of stakeholders in school meals can undergo changes every four years, when elections are held—changes that begin in the pre-election period, as the PNAE can be used as a political bargaining chip. With regard to family farmers, interviewees reported that in some locations “obstacles” were created for this group of stakeholders to participate in public procurement, such as requiring additional documents, lack of flexibility, lack of transparency in public procurement, and excessive bureaucracy. There were also reports of favoritism toward certain groups and the ease with which food industry representatives can access local governments. In general, there was a sense of frustration among stakeholders in some municipalities regarding the creation of laws and the lack of support from politicians to foster an environment conducive to compliance.

Finally, our respondents mentioned a resistance to change,i.e. the difficulties in accepting the food changes prescribed by the laws. For example, in the case of some cooks, although they receive menus adjusted to the laws, it was reported by the Social Control actors and nutritionists that they refuse to follow them. The cooks claim that students do not accept the new menu and that a lot of food is wasted. As for the students, it was reported by cooks and nutritionists that after the pandemic many returned to poor eating habits and reacted negatively to the changes made to school meals. Interviewees mentioned that many of the foods offered at schools are not part of students’ daily diet at home, with limited availability and variety of fruits and vegetables, and they therefore reject certain foods with which they are not familiar.

Another example related to resistance to change reported by Cecane, nutritionists and cooks, was that representatives of the food industry (especially in smaller municipalities) still try to persuade municipalities to buy their products. In addition, teachers, principals, and parents also raise questions about the menu changes and don’t help to control unhealthy foods brought by students; and many have biased opinions about the menu, based on their own eating habits. Parents reported concerns about students going hungry (due to low acceptance of the foods offered at schools) and at the same time show a lack of interest in the quality of what is being offered at school. Many of those interviewed linked the parents’ resistance to the implemented laws to the fact that eating foods high in sugar is part of the region’s culture, and there is widespread consumption of ultra-processed foods among families who currently eat more processed and ready-to-eat foods.

## Discussion

To understand the implementation of a complex program such as PNAE within a social, economic, and geographic context as diverse as Brazil, this study draws on Lipsky’s^17^ theory of street-level bureaucracy, which examines the role of frontline actors in implementing public programs, policies, and laws. According to Lipsky^17^, discretion is the mechanism through which these actors translate policies and regulations into practice under varying institutional and contextual conditions. As Lipsky^17^ argues, the central question is not whether street-level bureaucrats exercise discretion—since this is inevitable—but rather how they will use it.

It was observed that freedom and autonomy were highlighted by the actors as positive aspects of their work. According to the literature^17^, those are essential elements which are part of the concept of discretionary practices. Autonomy, when granted to SLB, can not only have a positive impact on the implementation of public policies and delivering public services, it can also lead to an overall feeling of self-efficacy and personal motivation to execute daily tasks in their routine^22^. In work environments, the sense of autonomy is linked to the ability to solve problems, decision making and job satisfaction^23^ (Bienefeld et al.,2025). In the context of PNAE, where policy needs to be adapted to different geographical realities and decisions need to be made locally, autonomy makes it easier for problems to be solved directly at the source from which they arise^24,25^.

However, in order for SLBs to use the autonomy granted in a more effective manner, they are required to develop professional knowledge and to gain support from other actors and organizations^26^. Our findings related to actors’ networks as a boost to the implementation of the PNAE’s laws are aligned to the published literature, which concludes that good communication and a network of actors are important to foster SLBs’ confidence in areas in which they lack full expertise, which can play a constructive role by shaping the direction and outcomes of collaborative governance efforts^27^. Professional training and skills development, as well as collaboration among different actors, have the potential to help street-level bureaucrats better navigate the challenges inherent in their roles of implementing policies, programs and laws in real and complex scenarios^26^.

Discretion, using the analogy of an elastic band, can be expanded to empower street-level bureaucrats to perform their duties, or conversely to restrict their ability to perform their duties adequately. For example, political commitment was identified as both a barrier and a driver for the local policy implementation, indicating the diverse outcomes of actor discretion among the political stakeholders. Other studies also have reported on how a lack of political will generates adverse consequences for compliance with regulations and policies^28,29^. Lipsky^17^ pointed out that the most common limitations in public services, such as financial constraints imposed on street-level bureaucrats, impact how they conduct their activities. In other words, street-level bureaucrats may need to make choices, such as prioritizing activities or groups of people, or even distancing themselves and disengaging from their duties^17,30^. This study reported examples such as the purchase of products of inferior nutritional quality and flavor, and the need to make fruit last longer by dividing it into several pieces to serve all the students. Other studies have found difficulties in fulfilling the nutritionist’s responsibilities, developing an appropriate menu, or prioritizing economic aspects^31–33^.

Another barrier found in this study that deserves attention is that related to the food system. Although much progress has been achieved since Law No. 11.947 (2009) was implemented with regards to local farmers’ supply and consumption of their products by students, there is still room for improvement. The way that food system transformation has occured since the 1940’s led to some consequences such as inequalities in agriculture, trade liberalisation, land being used for the needs of big corporations commodities that still have a big impact on small local farmers^34^. Furthermore, in Brazil there is a strong influence of agribusiness in the policy arena debate, and the government still subsidizes foreign investments to the detriment of smallholder farmers and local agribusiness^35^. Continuous efforts by all actors from PNAE and civil society are necessary to achieve more progress, because fighting for a sustainable food system is a battle that historically has shown that the odds are against population health.

Several implications are evident from the findings of this study. Firstly, the results highlight the importance of organizations that provide training and technical support to stakeholders directly involved in the implementation of PNAE regulations. Strengthening the technical knowledge of these actors and facilitating collaboration among them proved crucial for building autonomy and the ability to problem-solve in the face of challenges such as high labor demands, workforce shortages, and limited financial resources. These organizations also appear to hold a relatively independent perspective on the functioning of PNAE’s institutional structure, which allows them to identify weaknesses in program implementation and propose solutions more closely aligned with the program’s operational needs.

Secondly, the interrelationships among stakeholders appear to play a central role in PNAE implementation. Interviewees frequently emphasized that the formation of collaborative networks among actors helps distribute workloads, facilitates the exchange of knowledge, and supports the acceptance of school meals among students. Strengthening these networks through targeted training and institutional support may therefore represent an important strategy for improving program implementation. Such efforts should also include key actors within the school environment—such as principals and teachers—as well as individuals responsible for food practices in students’ households.

Thirdly, across the municipalities studied, drivers and barriers to implementation were present to varying degrees. Regardless of regional context, stakeholders reported making efforts to implement the regulations as effectively as possible. However, municipalities with limited fiscal capacity face particular challenges in sustaining the investments required to implement PNAE effectively. In such contexts, both family farmers and students may not fully benefit from the program as intended, suggesting that additional support mechanisms or policy adjustments may be necessary to address these structural disparities.

Finally, family farmers play a central role in the implementation of PNAE, functioning both as suppliers and as beneficiaries of the program. Since the enactment of Law No. 11.947 (Brasil, 2009), important advances have been made in incorporating family farm products into school menus. Nevertheless, the present study indicates that significant challenges remain, including limited access to information and the bureaucratic demands imposed on these stakeholders.

One potential limitation of this study relates to the selection of interview participants. In most municipalities, nutritionists helped identify additional interviewees, which may have introduced selection bias if participants were chosen based on proximity, affinity, or shared perspectives regarding PNAE. However, this potential limitation may have been partially mitigated by the inclusion of participants from external organizations, who had no direct political or professional ties to the municipalities studied.

At the same time, the study also presents important strengths, including the high participation rate among invited stakeholders and the inclusion of municipalities from all five geographic regions of Brazil, which contributed to the diversity of perspectives recorded and strengthened the overall quality and relevance of the data collected.

## Conclusion

This study investigated the drivers and barriers to local PNAE implementation across the country’s five regions, drawing on the perspectives of key program stakeholders. By examining diverse local contexts, the study documents important nuances and lessons regarding how these regulations are implemented in practice. The Street-Level Bureaucracy framework^17^ proved useful for understanding the day-to-day dynamics faced by PNAE stakeholders. Through discretionary practices, frontline stakeholders rely on factors such as professional autonomy, collaborative work, technical training, human and physical resources, and political commitment to implement program regulations. However, when barriers outweigh drivers, stakeholders reported adapting their practices in ways that may restrict program activities and potentially compromise the delivery of nutritionally adequate school meals.

Although the drivers and barriers identified were broadly similar across the municipalities studied, their magnitude varied according to local socioeconomic and institutional conditions. In municipalities facing greater economic vulnerability and fiscal constraints, additional support and targeted actions from the Federal Government may be necessary to strengthen program implementation and ensure that both students and family farmers fully benefit from PNAE.

Given the scale of PNAE and its growing role as a global reference for school feeding policies, the lessons identified in this study may also inform the implementation of similar programs in other countries. Strengthening technical support networks, improving coordination among stakeholders, and addressing structural resource constraints are likely to be critical for translating school feeding regulations into effective practice. Finally, future interventions can explore strategies for strengthening the engagement of school community stakeholders—such as teachers, principals, and parents—with PNAE. These actors may play a key role in supporting the implementation of program regulations and improving students’ acceptance of school meals.

## Data Availability

All data produced in the present study are available upon reasonable request to the authors.

